# Late Life Learning, Cognition, and Aging: A Longitudinal study of middle and older-aged adults in Beirut, Lebanon (3LC study)

**DOI:** 10.64898/2026.09.10.26362603

**Authors:** Adina Zeki Al Hazzouri, Abla Mehio Sibai, Darina Bassil, M Maria Glymour, Jennifer J Manly, Maya Abi Chahine, Lara Chehabeddine, Mira Bekdache, Martine Elbejjani

## Abstract

**Background:** Lebanon is undergoing substantial demographic and social change, with older adults comprising approximately 10% of the population. The Late Life Learning, Cognition, and Aging (3LC) study was established around the University for Seniors (UfS), a longstanding late-life learning program at the American University of Beirut. The cohort was designed to leverage variation in late-life learning exposure by including both UfS participants and community-dwelling adults without prior participation in the program. This provides a unique opportunity to investigate how late-life learning, lifecourse social and biological factors, and health behaviors relate to cognitive aging, dementia risk, mental health, and resilience in Lebanon.

**Methods:** Participants aged 50 years and older are recruited from the UfS program and the surrounding community in Lebanon. Baseline assessments include an interviewer-administered questionnaire, standardized anthropometric and sensory assessments, and blood collection for biomarker measurement and biobanking. The questionnaire captures health, psychosocial, cognitive, social engagement, and lifecourse factors, including exposure to war, conflict, and socioeconomic adversity. Longitudinal follow-up includes planned phone-based and in-person assessments at approximately two-year intervals.

**Results:** The cohort comprised 1,871 participants, including 536 UfS participants (350 previously enrolled and 186 newly enrolled) and 1,335 community participants, of whom 1,041 were age-, sex-, and education-matched to UfS participants and 294 were additionally recruited without matching. The mean age was 65.6 years (SD=9.0), and 70.1% of participants were women. Blood samples were obtained from 1,541 participants (82.3%).

**Conclusion:** The 3LC study provides a unique platform to examine how late-life learning and lifecourse factors shape cognitive aging, well-being, and dementia risk in Lebanon. By integrating social, psychosocial, health, and biological measures, the cohort offers opportunities to identify modifiable determinants of cognitive aging and inform dementia prevention in Lebanon and other under-resourced settings.

## 1. INTRODUCTION

Population aging is one of the most significant demographic changes of the twenty-first century, with low-and middle-income countries (LMICs) projected to be home to more than two-thirds of the global older population by 2050^1^. As life expectancy increases, the number of people living with dementia is projected to exceed 139 million by 2050, underscoring its growing global public health burden^2^. It is currently estimated that over 60% of people with dementia live in LMICs, a proportion expected to exceed 70% within the next three decades^3,4^. Yet, many LMICs remain under-equipped to respond to population aging due to limited resources, constrained research infrastructure, and under-resourced health systems^5^.

Projections indicate that the Middle East and North Africa (MENA) region will face the steepest rise in dementia cases^2^. In parallel, population-based evidence on cognitive aging and dementia risk remains scarce, reflecting limited epidemiological and longitudinal research capacity across the region. Lebanon, a small LMIC on the eastern Mediterranean coast, represents a particularly important setting for investigating cognitive aging and dementia. It has one of the highest proportions of adults aged 65 years and older (10.4%) and is among the fastest-aging countries in the region^6,7^. This demographic transition is unfolding amid prolonged economic, political, and social instability^8^. Older adults in Lebanon have experienced repeated psychosocial adversities across the life course, spanning a 15-year civil war (1975–1990), recurrent armed conflicts, and multiple political and economic crises. More recently, these long-standing adversities were further compounded by a national economic collapse that began in 2019, ranked among the most severe economic crises historically, as well as the catastrophic Beirut Port Blast (2020), and renewed regional conflicts since 2024^9^. Despite this distinctive convergence of exposures, little is known about how these life course and recurrent adversities shape cognitive aging and dementia risk in Lebanon. Understanding these pathways is important for characterizing cognitive aging in Lebanon and broadening evidence on dementia risk and resilience in underrepresented populations^18^.

Furthermore, in Lebanon, as in many other LMICs, prospects for older adults to remain physically active, cognitively stimulated, socially connected, and engaged in their communities are limited^19,20^. Social connectedness, cognitive stimulation, and continued engagement are associated with better cognitive and health outcomes in older age, presenting important modifiable strategies for healthy aging^21^. Building and maintaining social capital in older age may be particularly important for buffering the cumulative effects of life course adversity amid increasing social isolation and recurrent economic and political disruptions^20^. In particular, there is growing interest in Late Life Learning (LLL) programs, which provide non-formal learning opportunities tailored to the interests and needs of older adults^20^, as a potential strategy to promote cognitive health, social engagement, and wellbeing in later life^22,23^. These typically low-cost and scalable programs are especially relevant to resource-constrained LMIC settings, where opportunities for healthy aging and social engagement are limited. In addition, while many determinants of cognitive aging, such as educational attainment are established earlier in life, LLL represents a window of opportunity in older age to modify accumulated risk and enhance cognitive resilience^21,24^. However, formal evaluations of LLL programs and empirical evidence of their cognitive and broader health benefits are largely lacking.

To address these interrelated gaps, we established the **L**ate **L**ife **L**earning, **C**ognition, and Aging (3LC) study, a longitudinal cohort of middle and older age adults in Beirut, Lebanon. The 3LC study examines whether LLL supports cognitive and overall wellbeing in older age while also investigating how psychosocial and biosocial pathways shape aging outcomes and dementia risk in a uniquely exposed and understudied population. By integrating social, economic, psychosocial, and biological measures, the study aims to identify modifiable determinants of cognitive aging and opportunities for dementia prevention in Lebanon and other LMIC settings. It also provides a resourceful platform for developing and validating innovative longitudinal methods, including telephone-based cognitive assessments and approaches to capture cumulative life course psychosocial adversity, while strengthening longitudinal research capacity and infrastructure to address current and emerging evidence gaps in Lebanon and other under-resourced settings.

## 2. METHODS

### 2.1 Study Design & Setting

The study was conducted in Lebanon, a country with an estimated population of approximately 5-6 million people, where population estimates are complicated by the absence of a recent census, and compounded by substantial and changing patterns of emigration, displacement, and long-standing refugee populations^25^. Lebanon is highly urbanized, with around 45% living in major urban and peri-urban areas, including Administrative Beirut and surrounding governorates. The country has also undergone substantial demographic transition, with older adults currently comprising 10-12% of the population, among the highest and fastest aging proportions in the region^7,8^.

Within this setting, the 3LC study was established around the University for Seniors (UfS), a long-standing LLL program at the American University of Beirut^26^. The UfS is the only LLL program in the region, recognized by RAND-WHO as one of the most innovative community-based programs in LMICs. Established in 2010, UfS provides educational, cultural, and social activities for adults aged 50 years and older and represents a unique platform for studying the role of LLL and social engagement in healthy aging. The 3LC study leverages the program’s long history and diverse LLL exposure patterns and includes UfS participants and community participants without prior LLL exposure. This enables investigation of LLL in relation to cognitive and health outcomes, and the role of broader life course social and biological factors in shaping cognitive aging, dementia risk and resilience.

### 2.2 Study Population & Eligibility

Launched in January 2025, 3LC is a community-based cohort of 2000 adults ages 50 and over residing in Beirut, the capital of Lebanon (mean age= 65.5 years (SD=9) and 30.3% men). The cohort includes 536 participants enrolled in the UfS LLL program and 1,331 community participants without LLL exposure. Participants completed a comprehensive baseline assessment, with planned consecutive phone-based, and in-person follow-up waves at approximately two-year intervals. **Figure 1** provides an overview of the study timeline.

**Figure 1.**
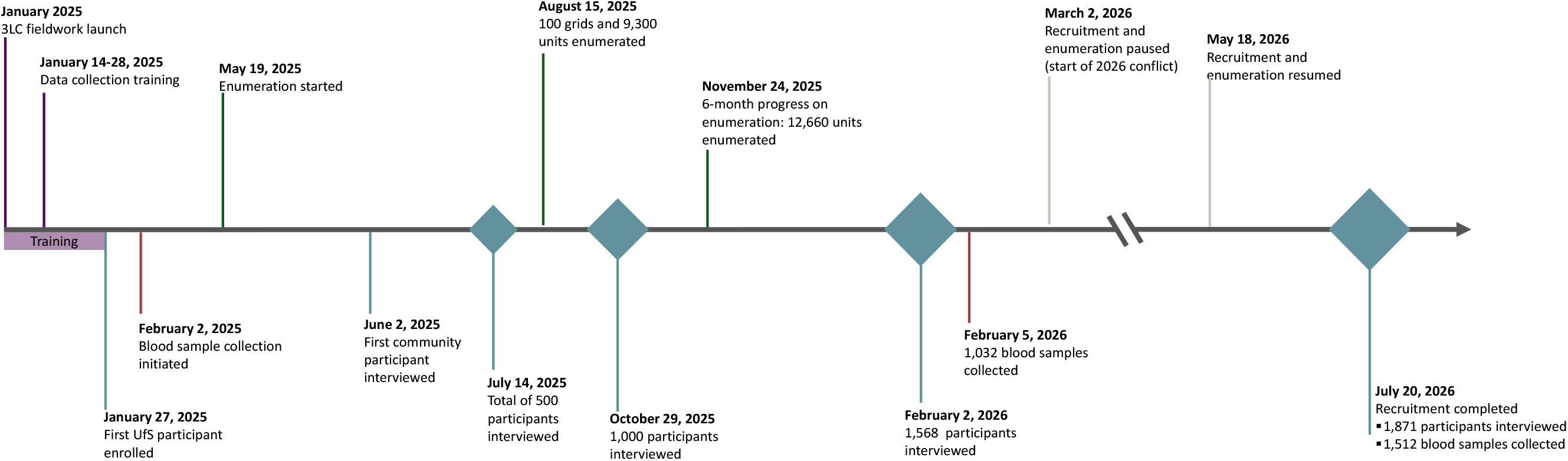
Study timeline, Late Life Learning, Cognition and Aging Study (3LC), Beirut, Lebanon, 2025-2026

Participants were eligible if they were ≥50 years old, residing in Beirut (for at least six months of the year), completed at least primary level education (i.e. grade 5), were able to leave their home independently or with assistance, and were able to complete the study questionnaire in Arabic. These criteria, especially education and mobility, were selected to enhance comparability between UfS participants and their community counterparts. Inclusion of participants aged 50 and older was designed to capture cognitive and health trajectories from midlife through older age, including transitions from normal cognitive aging to cognitive impairment and dementia.

### 2.3 Study Recruitment & Sampling

The 3LC cohort was assembled using two complementary sampling strategies that leveraged the UfS and optimized community-based recruitment in the absence of high-quality population sampling frames. The primary sampling strategy was designed around the LLL exposure and the objective of conducting LLL program evaluation and included three groups: (1) previously enrolled UfS participants, (2) newly enrolled UfS participants, and (3) age-, sex-, and education-matched community participants without exposure to LLL (**Figure 2**). A supplemental extended recruitment strategy was implemented to enroll eligible community participants who did not meet matching criteria, expanding the cohort for broader investigation of life course determinants of cognitive aging and related health outcomes.

**Figure 2.**
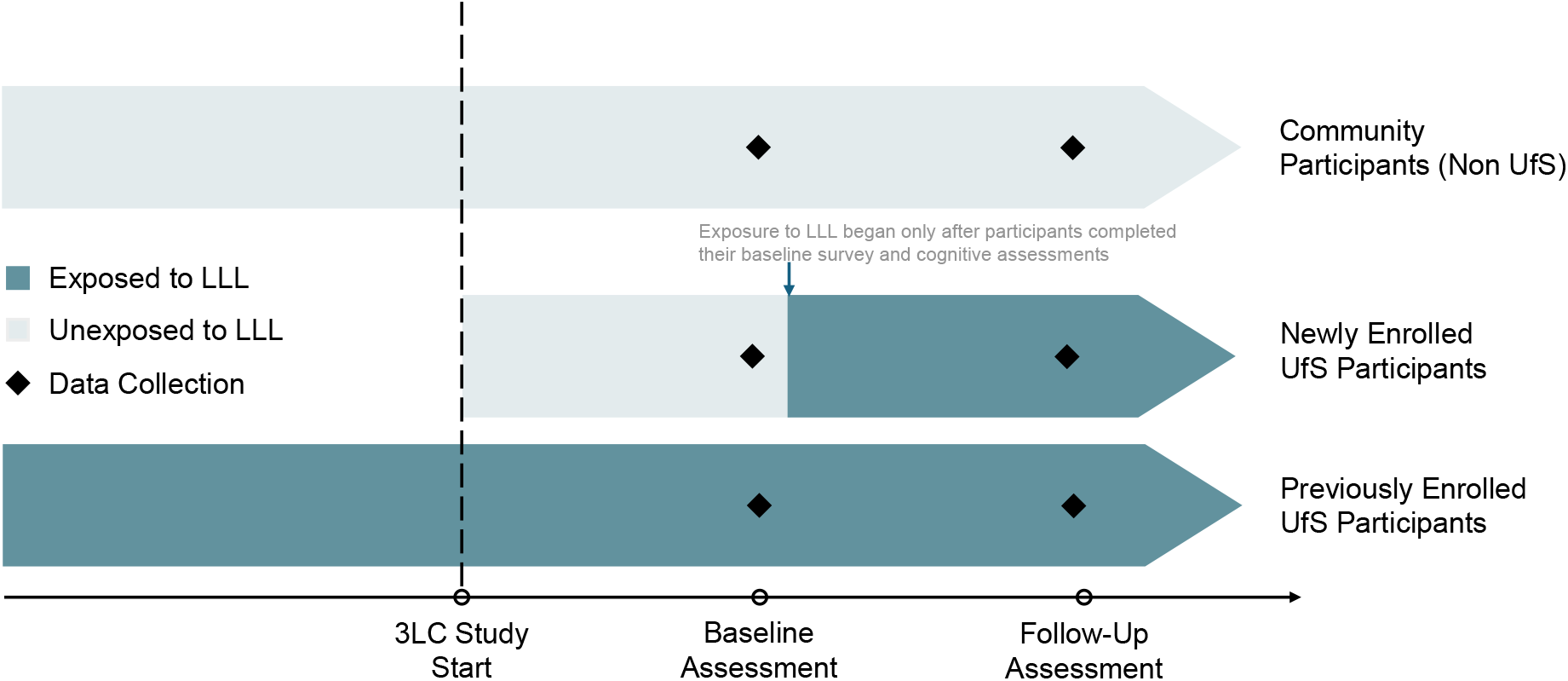
Illustration of the study design and exposure arms, Late Life Learning, Cognition and Aging Study (3LC), Beirut, Lebanon, 2025-202

#### 2.3.1 Sampling Strategy 1: Exposure-based Sampling

Because UfS has been running for over 16 years, the primary exposure-based sampling strategy incorporated both previously enrolled participants with varying durations of prior LLL exposure and newly enrolled participants who had not yet initiated the program at time of study baseline assessments.

Previously enrolled UfS participants (n=350) provided an opportunity to investigate associations of longer and real-world participation in LLL programs. However, because their LLL participation preceded study enrollment, pre-exposure measures of cognitive function, potential confounders, and determinants of participation could not be collected. To address this, additional recruitment of newly enrolled participants (n=186), who initiated LLL only after baseline assessments were completed, ensuring that we have measures on cognition and predictors of UfS participation prior to LLL exposure. Matched community participants without prior LLL exposure served as the comparison group. Together, these three groups support investigation of both long-term associations and prospective cognitive and health changes associated with LLL participation (**Figure 2**).

##### 2.3.1.1 Sampling strategies of previously enrolled UfS participants

Previously enrolled UfS participants were identified through administrative records of individuals aged ≥50 years who had participated in at least one term since 2017 (n=807). UfS personnel initially contacted eligible members to obtain permission to share their contact information with the study team. Those who agreed were subsequently contacted by phone and invited to participate, resulting in the recruitment of 350 prior UfS members.

##### 2.3.1.2 Sampling of newly enrolled UfS participants

The UfS program runs two terms annually, with approximately 50 new registrants per term. Individuals newly enrolled in Spring 2025, Fall 2025, and Spring 2026 terms (N=303) were invited to join the study using the same recruitment procedures, yielding 186 participants with baseline assessments completed before the UfS term began, ensuring measures were collected prior to LLL exposure.

##### 2.3.1.3 Sampling of non-UfS community participants

Residential addresses of previously and newly enrolled UfS participants were obtained to support recruitment of matched community participants. This strategy allowed identification of sampling grids, thus overcoming the lack of reliable sampling frames in Lebanon. Using ArcGIS Pro, a GIS-based sampling frame was developed to support field enumeration across administrative Beirut. Districts and neighborhoods were partitioned to 100*100m, 200*200m or 400*400m grids based on population density. Grids containing UfS participants’ addresses were designated as anchor grids. Of the 222 UfS anchor grids identified, 25 located in high-security neighborhoods were excluded, leaving 197 grids eligible for enumeration. Fieldwork was initiated in four grids, purposely chosen for proximity to AUB, to facilitate initial field implementation. Remaining eligible anchor grids were randomly ordered for enumeration. Each selected anchor grid was mapped together with its adjacent grids (8 surrounding grids per anchor grid), and up to two neighboring grids were randomly selected and assigned as extension grids to enhance sampling efficiency and neighborhood coverage. More broadly, the selection strategy was centered on grids containing known UfS addresses and then randomly expanded into eligible adjacent grids, thereby balancing recruitment efficiency with geographic coverage (**Figure 3**).

**Figure 3.**
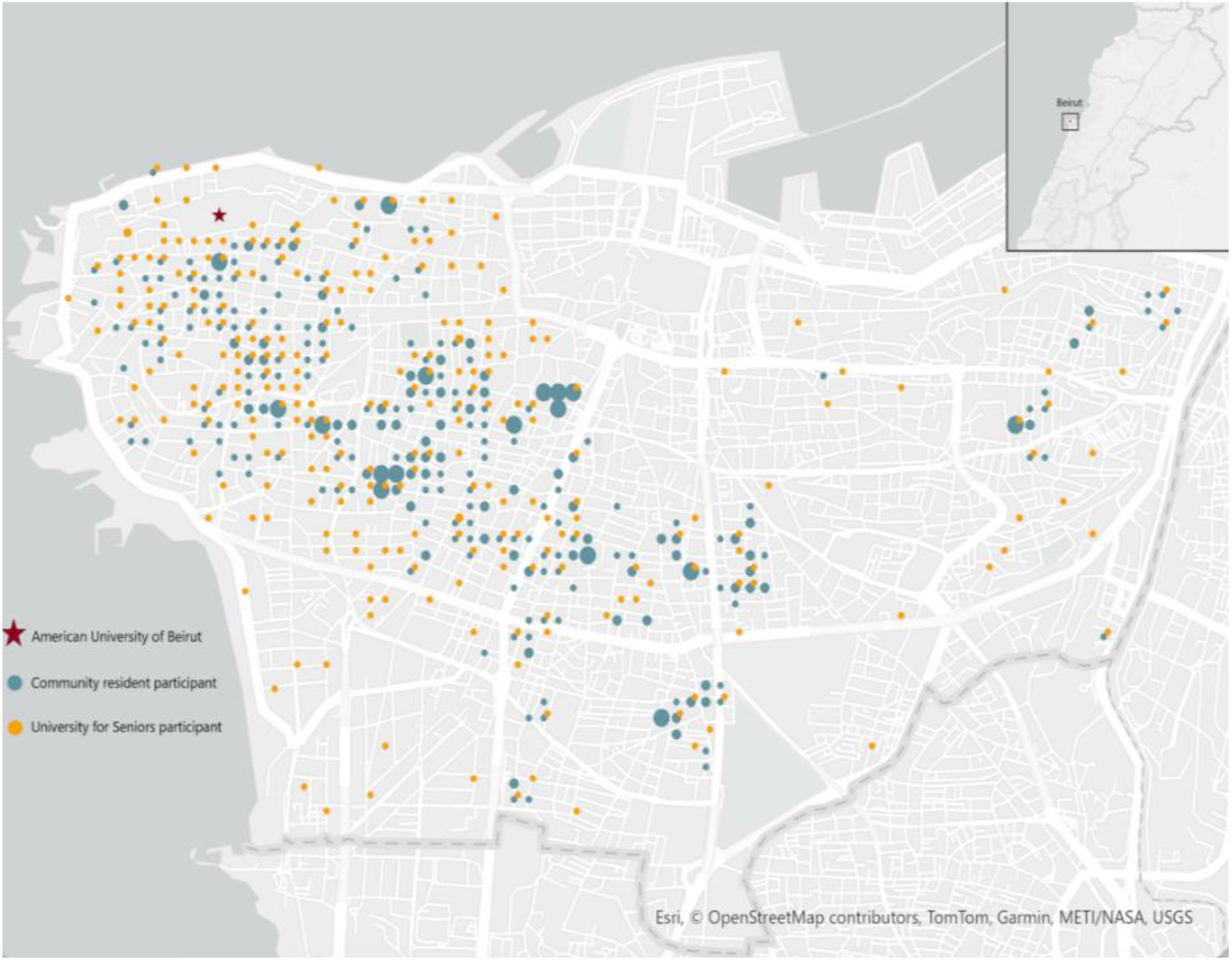
Mapping of recruited participants in the Late Life Learning, Cognition and Aging Study (3LC), Beirut, Lebanon, 2025-2026.

Within each selected grid, fieldworkers systematically enumerated households. Community participants meeting the study general eligibility criteria (age ≥50; education ≥grade 5; Arabic-speaking, and ability to leave the home) and with no prior participation in any LLL program were considered for recruitment. To improve sampling efficiency, up to two eligible individuals could be recruited from the same household, consistent with practices used in large-scale epidemiological cohort studies. Overall, 272 participant pairs (544 total participants) were recruited from the same household.

###### Matched community participants

Eligible community participants were matched to UfS participants according to age (within ±3 years), sex, and educational attainment (some university education or higher vs. no university-level education). Because UfS participants were more likely to be women and of higher educational attainment than the general population, matching at the design stage will ensure that we have enough numbers at the intersection of these potential confounders and hence a better control for confounding. UfS participants with higher education were matched 1:1 to community participants, whereas those with lower education were matched 1:8, to increase representation of participants with lower educational attainment.

#### 2.3.2 Sampling Strategy 2: Community Enrichment Sampling

To optimize recruitment efficiency and enhance cohort diversity, eligible community residents identified during household enumeration, but who were not matches to UfS participants were also invited to participate in the study, resulting in the recruitment of 294 additional community participants. In addition to increasing the total sample size, their inclusion diversified the sample for broader analyses addressing risk and protective factors of cognitive aging and related outcomes that do not require matching on LLL exposure.

The 3LC study was approved by the Institutional Review Board (IRB) at the American University of Beirut (BIO-2023-0405) and Columbia University (AAAV3042(M00Y01)). All participants provided written informed consent following a brief decision-making competency test to ensure proper understanding of study protocols and ability to consent.

### 2.4 Data Collection Procedures

#### 2.4.1 Field Training & Quality Assurance

Prior to data collection, fieldworkers received specific training on study protocols and standardized procedures. Data collectors completed an intensive nine-day training program covering informed consent procedures, digital data collection, administration and scoring of cognitive and psychosocial measures, anthropometric and sensory assessments, and documentation and reporting of field challenges. Following training, all data collectors were required to demonstrate competency through simulation-based practice assessment, didactic session, role-playing exercises, and cognitive score calibration exercises.

Enumerators completed a separate three-day training focused on household enumeration, participant screening and recruitment, and safety procedures; field supervisors received additional training on systematic building listing procedures and coordination of field teams. To reinforce study protocols adherence and address challenges encountered during fieldwork, refresher trainings were conducted approximately every 1.5 months.

Field activities were closely supervised throughout recruitment and data collection. A multi-level quality assurance strategy included routine analytical data checks to identify patterns of missingness, inconsistencies, and implausible responses, as well as random field observations and review of audio-recordings of interviews to assess adherence to standardized administration and scoring procedures. Data quality reports were routinely generated and used to provide timely individualized feedback to data collectors and to guide refresher training when needed, thereby ensuring continued standardization across the field team.

Prior to field implementation, the study questionnaire and assessment procedures were pilot tested for clarity, comprehension, flow, and estimated administration time. A structured field pilot was subsequently conducted with a subset of UfS participants, and feedback from both participants and data collectors was used to refine study procedures before baseline data collection.

#### 2.4.2 Baseline Assessment and Data Collection

Baseline data collection was conducted between January 2025 and August 2026, primarily in participants’ homes, with assessments at AUB available according to participant preference. Assessments were conducted in Arabic by 15 trained data collectors using the surveyCTO and lasted approximately 2.5 hours.

Baseline data collection included three main components: an interviewer-administered questionnaire, standardized anthropometric and sensory health assessments, and blood collection for biomarker measurement and biobanking.

The study questionnaire provided a comprehensive assessment of demographic and socioeconomic characteristics, physical and mental health, psychosocial and social factors, LLL and broader social engagement, cognitive function, and life course exposures including exposure to war, conflict, and socioeconomic adversity. Questionnaire design prioritized instruments and measures previously validated or used in Lebanon. Where suitable measures were unavailable, items were adapted or developed drawing on prior literature and measures used across established international aging cohorts, particularly the Health and Retirement Study (HRS) network in LMIC settings. Measures without existing Arabic-language versions were translated and reviewed for linguistic and cultural appropriateness. The cognitive battery was developed in alignment with the Harmonized Cognitive Assessment Protocol (HCAP), supporting harmonization and cross-national comparisons. **Table 1** summarizes measures collected in the 3LC baseline assessment.

**Table 1.**
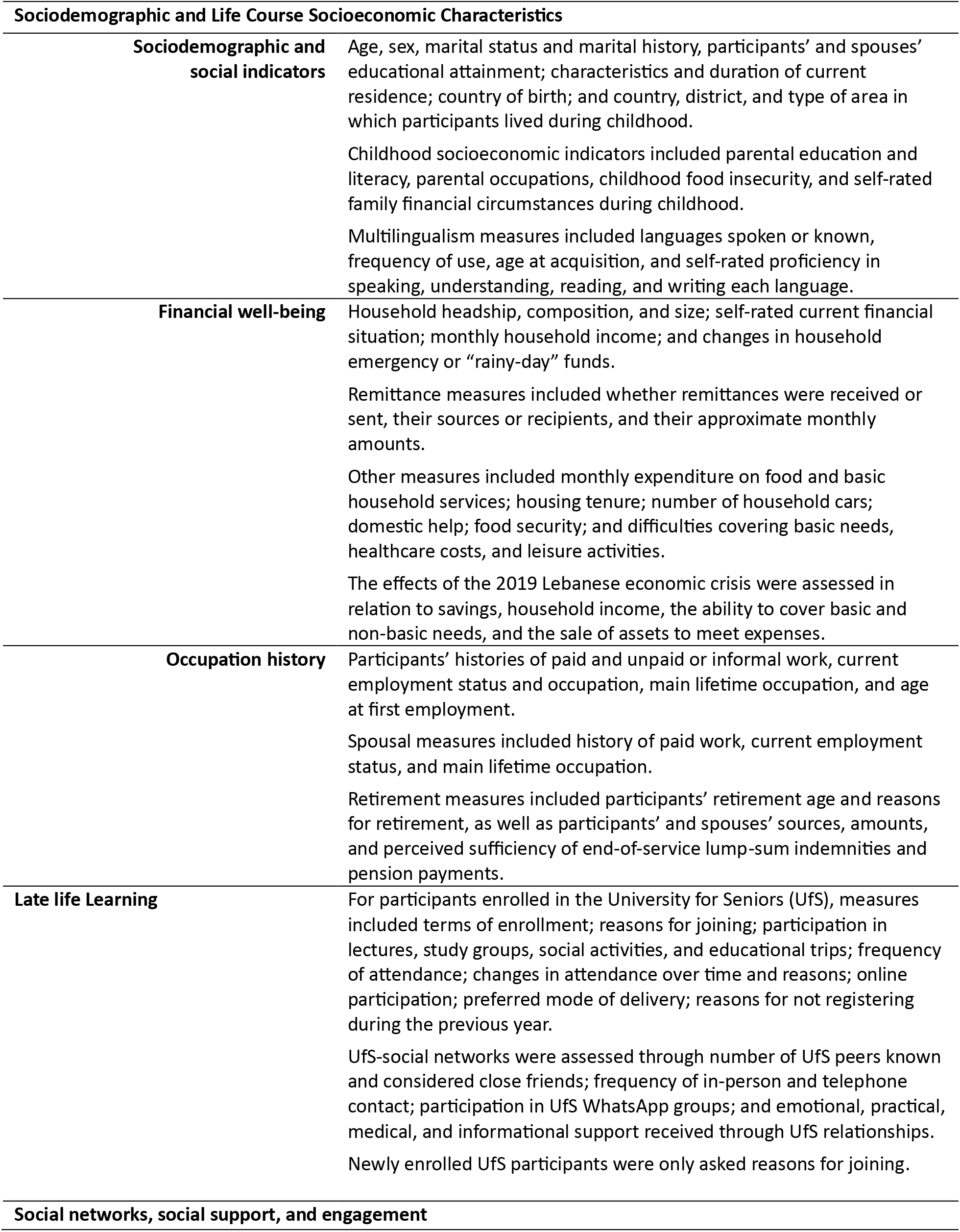
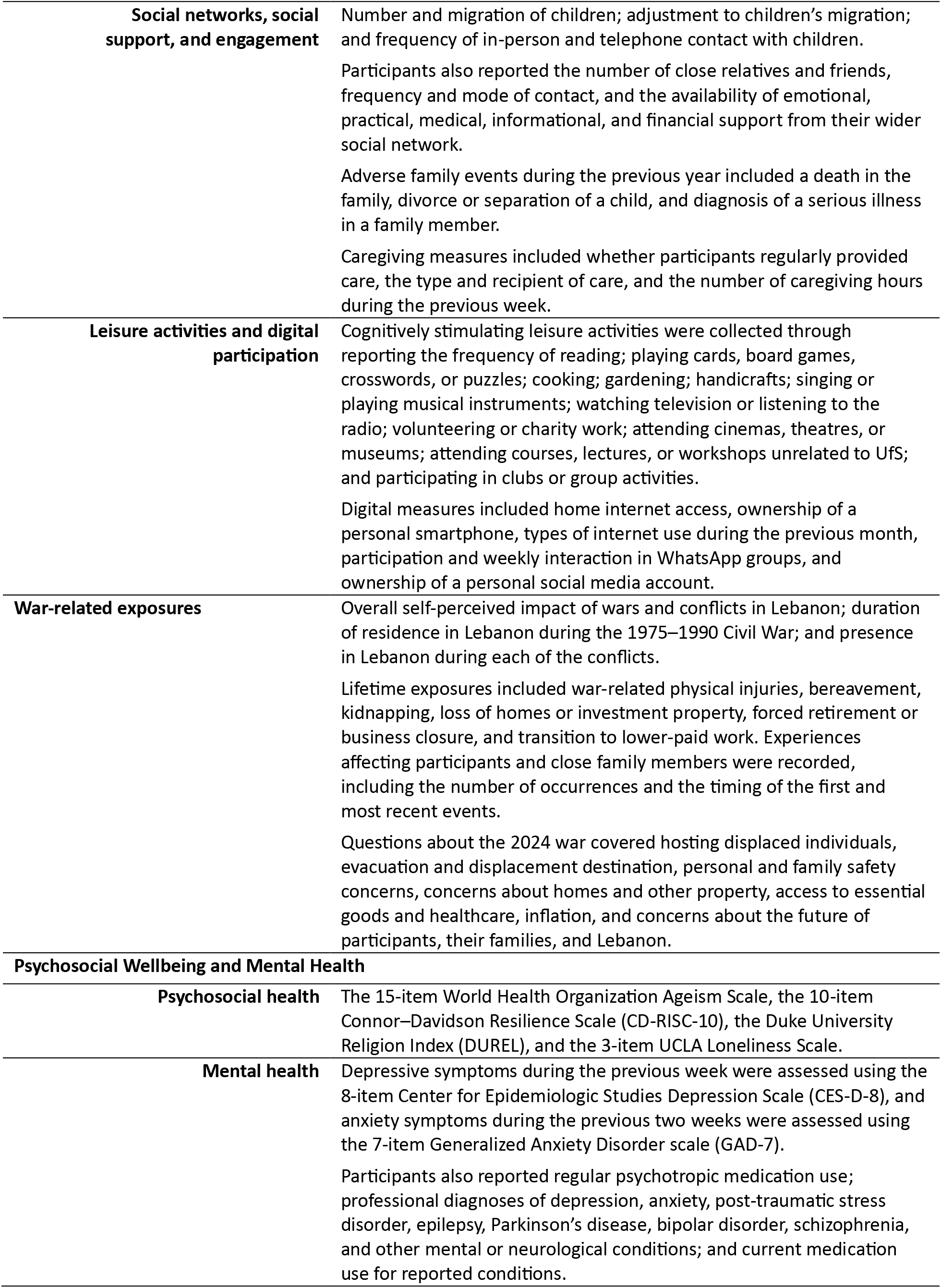
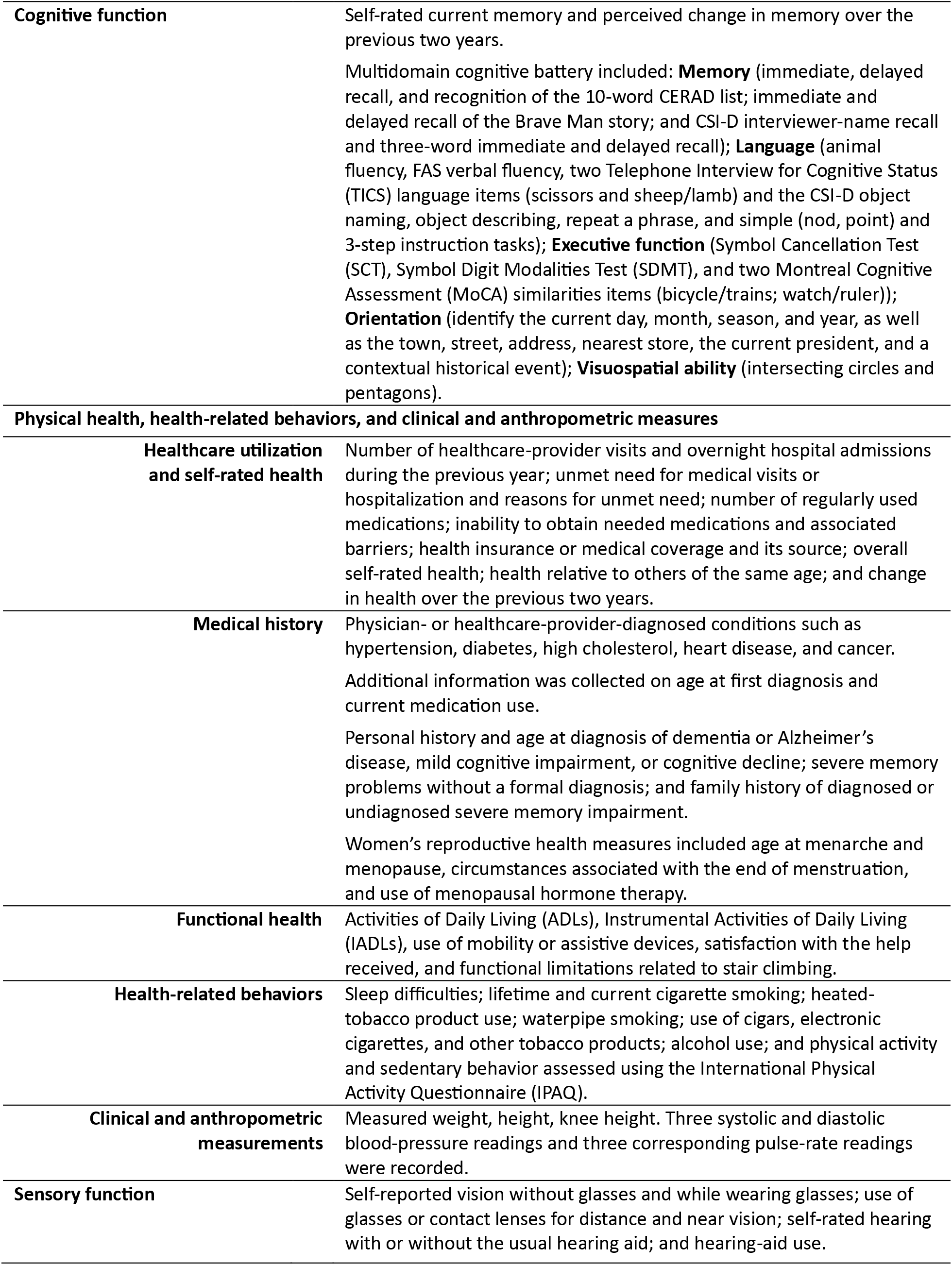
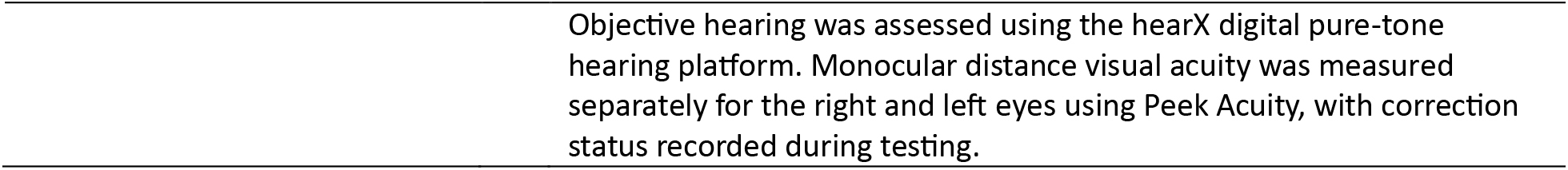
Data collected during in-person interviews, Late Life Learning, Cognition and Aging Study (3LC), Beirut, Lebanon, 2025-2026.

### 2.5 Measures

#### 2.5.1 Sociodemographic and Life Course Socioeconomic Characteristics

##### 2.5.1.1 Sociodemographic and social indicators

Measures included age, sex, marital status, education, place of birth, residential mobility, and childhood socioeconomic indicators. Given the high prevalence of multilingualism in Lebanon and its potential relevance to cognitive aging, the questionnaire included measures of languages spoken and frequency of use, age at language acquisition, and self-reported language proficiency.

##### 2.5.1.2 Financial Well-being

Measures included household income, remittances, financial assistance, household expenditures, food insecurity, and self-reported financial difficulties. The financial wellbeing section also included questions on how the 2019 financial collapse in Lebanon affected household income, personal savings, and ability to meet financial needs.

##### 2.5.1.3 Occupation History

This module captured participants’ and spouses’ employment status and occupational histories, including lifetime occupation, unpaid or informal work, retirement, and pensions and end-of-service benefits and sufficiency.

#### 2.5.2 Late Life Learning

UfS offers several options of engagement, ranging from lectures (topics such as history, science, health, and arts) to more focused study-groups aimed at learning specific skills (such as technology use, gardening, painting, or learning new languages). UfS members have access to all lectures offered during the term (19-25 lectures per term) and may also enroll in up to two study groups per term. Lectures and study groups are delivered on a voluntary basis by AUB and other university faculty, community experts, and UfS members. The program also provides educational and cultural activities, including museum visits and cultural trips (4-5 per term) and opportunities for social engagement through organized social events and gatherings (6-8 social activities per term). Table 2 provides the UfS Spring 2025 schedule as a sample of program offerings. Before the COVID-19 pandemic, all offerings were delivered in person; currently, approximately 40-60% are delivered online.

**Table 2.**
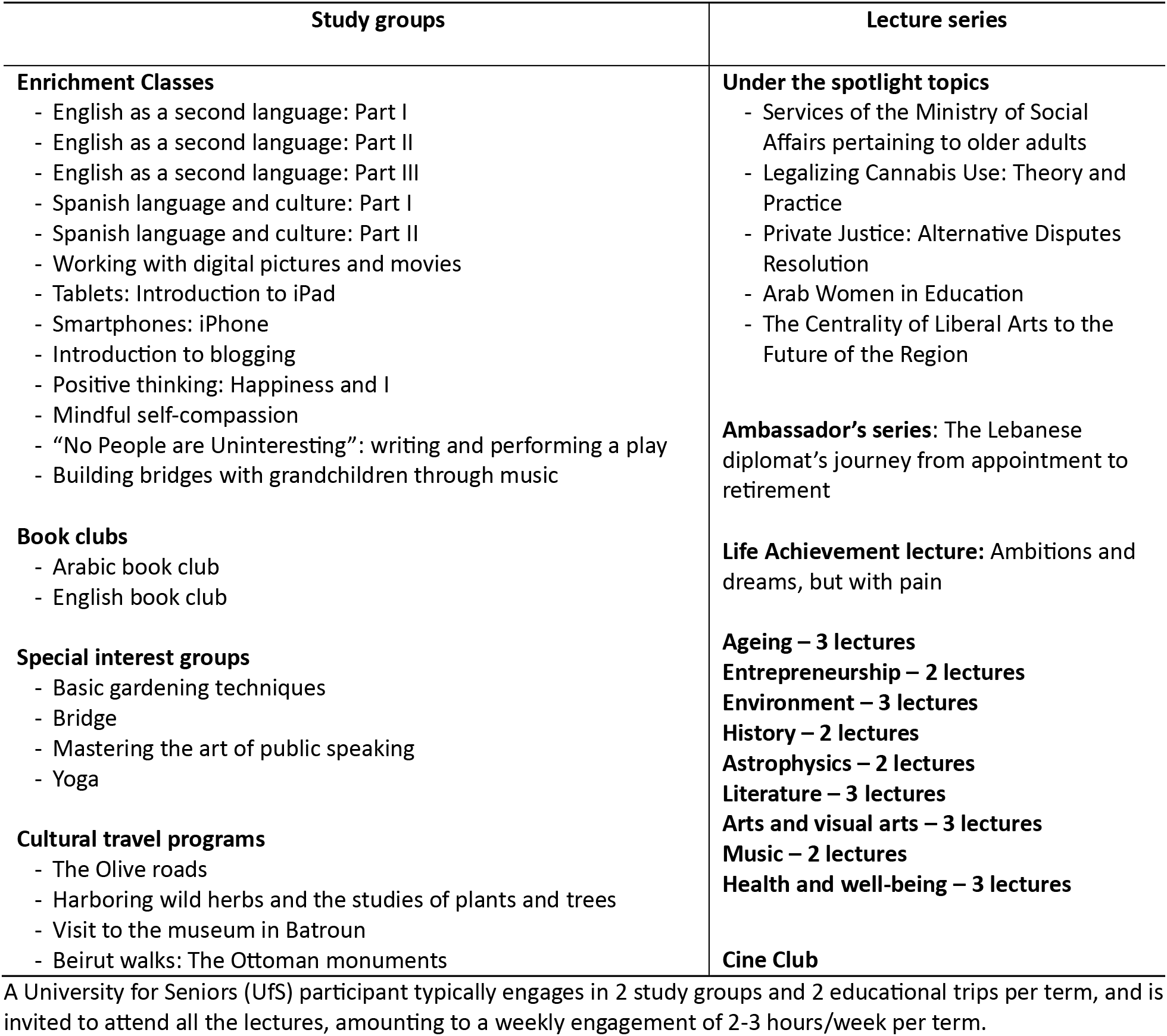
Sample program for the University for Seniors Spring term 2025, American University of Beirut.

LLL exposure was characterized across multiple dimensions, including duration and continuity of participation, cumulative exposure, type and frequency of LLL participation and differentiation between predominantly educational/cognitive and social forms of engagement. Because participation in UfS may influence social networks and support, participants also reported social connections developed through the program, including number of UfS peers and friends, frequency of contact, and the extent to which they could rely on these relationships for support.

#### 2.5.3 Social Networks, Social Support and Engagement

Measures covered family and social networks, including number of children, relatives, and friends, and frequency and mode of contact, and emotional, practical, medical, financial, and informational support. Given the prevalence of adult emigration in Lebanon, information was collected on children living abroad and participants’ coping with children’s migration. Caregiving activities, including type and frequency of care provided to others, were also assessed. Participants reported the frequency of engagement in a broad range of cognitively stimulating activities, including reading, playing cards and boardgames, volunteering, and participating in clubs or group activities. Together these measures allowed a better characterization of general cognitive function and social engagement as well as those developed through LLL participation.

The questionnaire also captured indicators of digital access and connectedness, an increasingly important component of social engagement and informational resources.

#### 2.5.4 Life course War-Related Exposures

Given Lebanon’s history of recurrent wars and armed conflict, lifetime war exposure was assessed across two broad domains: bodily harms (injuries, kidnappings and death) and material losses (loss of homes/property/businesses and employment) experienced personally by participants or close relatives. Information was also collected on frequency of exposure and timing of first and most recent events in each domain. Participants additionally reported the overall self-perceived impact of wars on their lives and whether they were residing in Lebanon during major periods of wars and violence. The questionnaire also captured experiences related to the more recent 2024 war, including personal and family displacement, safety concerns, financial difficulties, and access to essential needs. Together, these measures enable examination of consequences of recurrent war-related adversity across the life course, including recent exposures.

#### 2.5.5 Psychosocial Wellbeing and Mental Health

##### 2.5.5.1 Psychosocial Health

Participants were administered a set of validated psychosocial questionnaires. These included ageism using the 15-item World Health Organization (WHO) Ageism Scale^27^; personal resilience measured using the 10-item Connor–Davidson Resilience Scale (CD-RISC-10)^28,29^; and loneliness measured using the 3-item UCLA Loneliness Scale^30^. Religiosity and religious involvement were assessed using the Duke University Religion Index (DUREL)^31^.

##### 2.5.5.2 Mental Health

Standardized questionnaires of current mental health symptoms, including the Center for Epidemiologic Studies Depression Scale (CES-D)^32^ and the Generalized Anxiety Disorder 7-item scale (GAD-7)^33,34^ were administered. Participants reported on use of psychotropic medications and whether they had received professional diagnoses of depression, anxiety, post-traumatic stress disorder, epilepsy, Parkinson’s disease, bipolar disorder, and schizophrenia.

#### 2.5.6 Cognitive Function

##### 2.5.6.1 In-person Cognitive Battery

The 3LC study administered a comprehensive multidomain cognitive battery assessing memory, executive function, language, orientation, and visuospatial ability. The battery was based on the Harmonized Cognitive Assessment protocol (HCAP)^35^ and built on its construct-centered adaptation and implementation in the Lebanese Study of Healthy Aging (LSAHA)^36^, with minor additions and modifications, facilitating harmonization and comparisons across the growing global network of HCAP-based studies.

Memory was assessed using three trials of the 10-word CERAD immediate, followed by delayed word recall and recognition tests (CERAD)^37^, the Brave Man immediate and delayed story recall tests, and the Community Screening Instrument for Dementia (CSI-D) delayed name recall and 3-word immediate and delayed recall^38–40^. Executive function was evaluated using the Symbol Cancellation Test (SCT)^41^, and the Symbol Digit Modalities Test (SDMT)^42^, and two Montreal Cognitive Assessment (MoCA) similarities items (bicycle/trains; watch/ruler)^43^. Language was assessed through the animal fluency test^44,45^, the FAS verbal fluency test^44,45^, two Telephone Interview for Cognitive Status (TICS) language items (scissors and sheep/lamb) and the CSI-D object naming, object describing, repeat a phrase, and simple (nod, point) and 3-step instruction tasks^46^. Orientation was assessed by asking participants to identify the current day, month, season, and year, as well as the town, street, address, nearest store, the current president, and a contextual historical event^46^. Visuospatial ability was assessed using the intersecting circles and pentagons^46^. Self-rated memory was also assessed, including participants’ perception of their current memory status and changes in memory over the past two years).

##### 2.5.6.2 Phone-based Cognitive Battery

Recognizing the need for scalable and accessible approaches to longitudinal cognitive assessment in Lebanon and other resource-constrained settings, a phone-based cognitive assessment was developed and administered to a subsample of 427 participants, 4-6 weeks following the in-person baseline assessment. Assessments were administered by the same data collectors following training on phone-based cognitive assessment procedures. The phone-based assessment closely mirrored the in-person battery except for tests requiring visual administration which could not be administered by phone (the SCT, SDMT, and the two visuospatial items). This substudy allows the evaluation of the feasibility and comparability of phone-based and in-person cognitive assessments to inform and facilitate longitudinal follow-up strategies.

#### 2.5.7 Physical Health, Health-Related Behaviors, and Clinical and Anthropometric Measures

Participants were asked to self-rate their health and report whether they had ever been diagnosed by a physician with a range of medical conditions, including hypertension, high cholesterol, diabetes, heart disease, stroke, cancer, and respiratory conditions. Information on age of diagnosis and medication use was also collected. Participants also reported any dementia and cognitive impairment diagnoses and family history of dementia, cognitive impairment, or severe memory problems. Additional measures captured falls and fractures history, pain, unintentional weight loss, self-reported vision and hearing impairment and use of aid devices, and for women, reproductive health history. Physical functioning was assessed using Activities of Daily Living (ADL)^47^ and Instrumental Activities of Daily Living (IADL) measures^48^. Information on healthcare utilization, insurance coverage, and barriers to healthcare access was also collected **(Table 2).**

Health-related behaviors included sleep, cigarette and waterpipe smoking, other tobacco use, and alcohol consumption. Physical activity was assessed using a short version of the International Physical Activity Questionnaire (IPAQ), including type and duration of work, exercise (vigorous, moderate), and sedentary activity over the past week^49^ **(Table 2).**

Anthropometric and clinical measures included standardized measurements of weight, height, knee height, heart rate, and systolic and diastolic blood pressure were collected. Blood pressure was measured after a period of rest, with three readings obtained and averaged. All measurement equipment was routinely calibrated to ensure standardization, accuracy, and consistency across participants **(Table 3).**

**Table 3.** Anthropometry, physical performance and point of care measures, procedures for data collection and threshold values, Late Life Learning, Cognition and Aging Study (3LC), Beirut, Lebanon, 2025-2026.

| Anthropometry | Equipment | Field Procedures | Thresholds |
| --- | --- | --- | --- |
| Height | SECA® Stadiometer 213 | Measured in centimeters to one decimal place using a portable stadiometer, with the participant standing upright, barefoot, and head positioned in the Frankfort horizontal plane. | None |
| Weight | SECA® 803 Digital Flat Scales | Measured in kilograms to one decimal place using a calibrated digital scale, with the participant standing barefoot at the center of the scale and weight evenly distributed on both feet. | None |
| Knee height | Measuring tape | Measured in centimeters to the nearest 0.5 cm using a non-stretchable measuring tape, with the participant seated, knee flexed at 90°, and the tape placed from the heel to the anterior surface of the thigh above the knee along a straight line. | None |
| Body Mass index (BMI) | | $\frac{\text{Weight in kilograms}}{(\text{Height in meters})^2}$ | Underweight < 18.5;<br>Normal 18.5-24.9;<br>Overweight 25-29.9;<br>Obese ≥ 30.0 |
| Blood pressure | Omron® M3 Comfort | Three measurements, 1 minute apart, pulse recorded. Final blood pressure: average of the three measurements. | Hypertension: systolic ≥ 140 mmHg |
| Hearing test | HearX® Heartest | Hearing assessed using headphones in a quiet setting, participants indicated when tones were heard to determine hearing threshold. | Reference Range (dB):<br>Normal hearing: ≤ 25<br>Mild Impairment: 26-40<br>Moderate Impairment: 41-60<br>Severe Impairment: ≥ 61 |
| Vision test | Peek vision® | Vision assessed at 2 meters using a tablet, participants covered one eye at a time and indicated the direction of the letter 'E'. | Reference Range (logMAR):<br>Normal Vision: ≤ 0.3<br>Mild Impairment: 0.4 – 0.47<br>Moderate Impairment: 0.48- 0.9<br>Severe Impairment: ≥ 1.0 |

#### 2.5.8 Sensory Function

Hearing was assessed using the HearX digital hearing health platform, a portable, tablet-based system for efficient and cost-effective standardized hearing assessment in accordance with international assessment standards^50^. Equipment included a Samsung Galaxy 0Tab A7 tablet with the HearX application installed, and Sennheiser HD 280 Pro headphones. Study team members received extensive training from consultants affiliated with the HearX Group and the Cochlear Center for Hearing and Public Health at Johns Hopkins, followed by standardized training for data collectors. Hearing assessment was not administered to participants with cochlear implants, dizziness, an active ear infection or drainage, or ear pain or discomfort. Assessments typically required approximately 5–10 minutes to complete.

Vision was assessed using Peek Acuity, a standalone validated application designed to screen for visual impairment and measure visual acuity^51^. Scores are reported in standard LogMAR. Assessments typically required one minute per eye.

#### 2.5.9 Blood and Biomarker Samples

The 3LC Study collected and bio-banked whole blood samples for investigations of clinical and aging-related biomarkers. Blood samples (25cc) were collected from consenting participants (n= 1625, 83.72%) by trained phlebotomists at the American University of Beirut Medical Center. Participants were instructed to fast for 10-12 hours prior to sample collection.

Samples were processed for blood and DNA storage at - 80°C and for immediate processing of routinely assayed clinical markers, such as glycosylated hemoglobin (HbA1C) and fasting blood glucose, lipid profile (total cholesterol, low-density lipoprotein (LDL), and high-density lipoprotein (HDL), & triglycerides), and inflammatory markers such as C-reactive protein (CRP). After baseline assessment completion, participants received feedback on point-of-care blood results, blood pressure, anthropometric, vision and hearing assessments. Participants with potentially clinically relevant findings were advised to seek appropriate medical follow-up.

## 3. RESULTS

### 3.1 Cohort Recruitment and Composition

Among individuals identified as eligible for participation, 1,110 UfS participants and 2,037 community residents were considered for enrollment. Of the eligible UfS members, 718 (64.7%) agreed to be contacted by the 3LC study team, of whom 536 (74.7%) enrolled and completed the baseline interview. Of the eligible community residents, 1,335 enrolled and completed the baseline interview, corresponding to an enrollment rate of 65.5%. Reasons for non-enrollment included inability to establish contact (16.6%), refusal to participate (65.2%), and other factors (**Figure 4**). Among enrolled community residents, 1,041 were matched to UfS participants and included in the primary cohort sample, while an additional 294 eligible but non-matched participants were retained as a supplemental sample.

**Figure 4.**
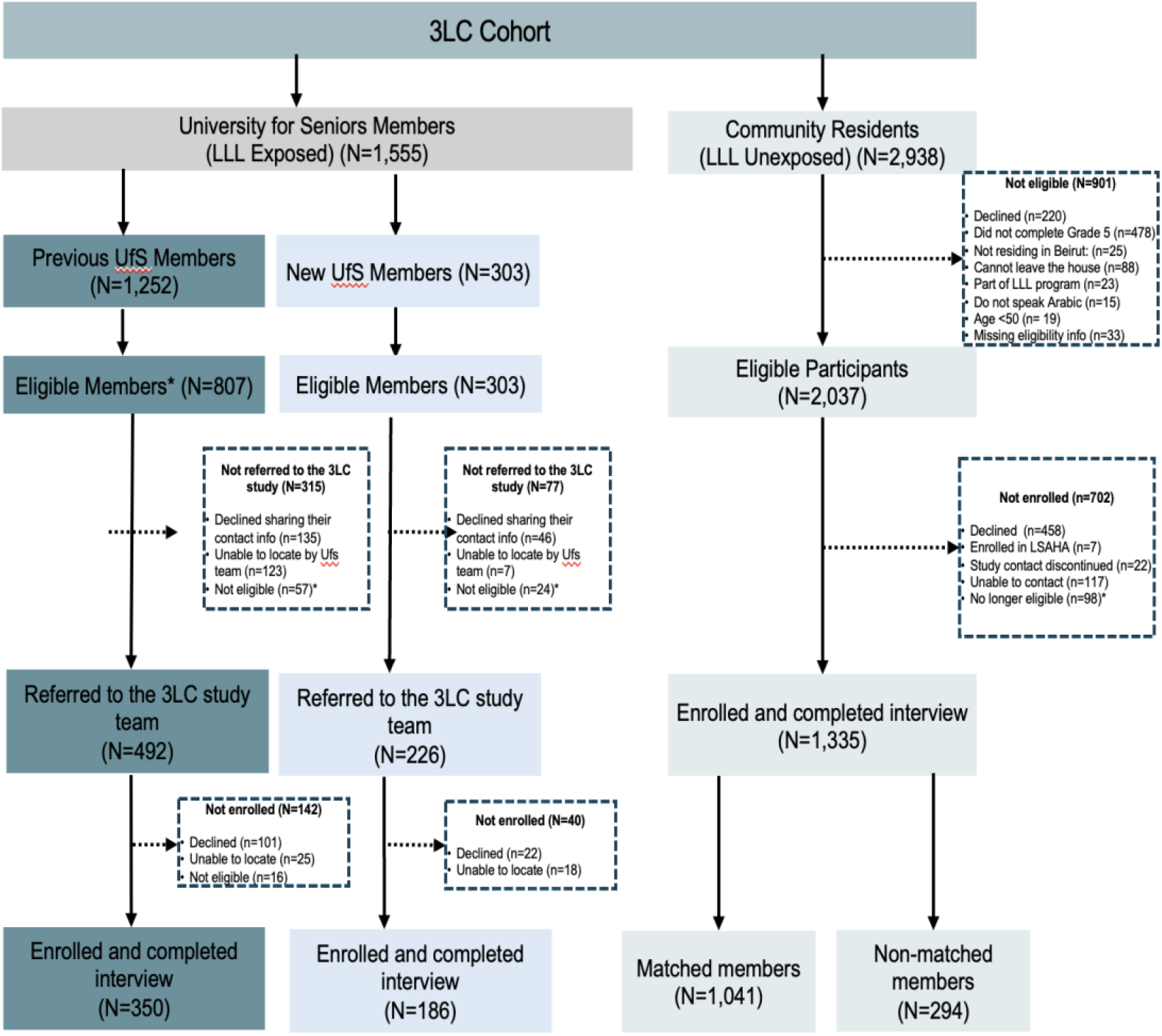
Study flowchart, Late Life Learning, Cognition and Aging Study (3LC), Beirut, Lebanon, 2025-2026

The final cohort consisted of 350 previously enrolled UfS members, 186 newly enrolled UfS members, and 1,335 community residents (**Figure 4**).

Completion of baseline assessment modules was high across the cohort, with minimal missing data across the different survey modules. Most modules had completion rates exceeding 99%, with the highest levels of missingness observed for anthropometric measures (1.8%) and hearing and vision assessments (2.4%)

Blood samples were obtained from 1,541 participants who consented to blood collection (82.3% of the full cohort). Consent rates were high and comparable across all recruitment groups, including 83.3% for newly enrolled UfS participants (n=155), 80.3% among previously enrolled UfS participants(n=281), 86.3% for matched community residents (n=898), and 89.1% among the supplemental unmatched community residents (n=262).

### 3.2 Baseline Characteristics of the 3LC cohort

The mean age in the full cohort was 65.6 years (SD=9.0), and the majority of participants were women (70.1%). Nearly two-thirds of participants were married (63.4%), while 18.3% were widowed. Educational attainment varied across recruitment groups, with 38.3% of participants having completed university education; higher levels of educational attainment were more common among UfS participants. Overall, 27% of participants were employed in full or part-time work at time of study enrollment.

Participants had a mean BMI of 29.07 kg/ m² (SD=5.67), with the majority classified as overweight (39.5%) or obese (37.0%). Self-rated health varied across groups, with most participants reporting good or fair health, although community residents generally reported poorer health compared with UfS participants. Cardiometabolic conditions were frequently self-reported including hypertension (48.5%), hypercholesterolemia (52.7%), and diabetes (23.5%). Smoking history was common, with 30.4% of participants reporting having smoked in their lifetime. Physical activity levels varied across the cohort, with 10.6% reporting vigorous activity and 56.7% reporting moderate activity. Depressive symptoms were also common, with a mean CESD-8 score of 8.69 (SD=6.04) and 44.9% of participants meeting criteria for elevated depressive symptoms based on the CESD-8 binary cut-off (≥9). Elevated depressive symptoms were more frequently observed among community residents compared with UfS participants.

Baseline characteristics of the 3LC cohort and individual recruitment subgroups are presented in **Table 4**.

**Table 4.** Baseline Demographics of the study sample, Late Life Learning, Cognition and Aging Study (3LC), Beirut, Lebanon, 2025-2026.

|  | **Total sample** | **Total matched sample** | **UfS- Previous** | **UfS- New** | **Community participant matched** | **Community participant Supplemental** |
| --- | --- | --- | --- | --- | --- | --- |
|  | **N = 1,871** | **N=1,577** | **N =350** | **N = 186** | **N =1,041** | **N = 294** |
| **Age, years, mean (SD)** | 65.6 ± 9.0 | 65.7 ± 8.9 | 69.4 ± 8.2 | 63.0 ± 8.1 | 64.9 ± 8.8 | 65.4 ± 9.9 |
| **Gender, n (%)** |  |  |  |  |  |  |
| Male | 560 (29.9%) | 278 (17.6%) | 59 (16.9%) | 45 (24.2%) | 174 (16.7%) | 282 (95.9%) |
| Female | 1311 (70.1%) | 1299 (82.4%) | 291 (83.1%) | 141 (75.8%) | 867 (83.3%) | 12 (4.1%) |
| **Marital Status, n (%)** |  |  |  |  |  |  |
| Married/partnered | 1186 (63.4%) | 948 (60.1%) | 213 (60.9%) | 127 (68.3%) | 608 (58.4%) | 238 (81.0%) |
| Never married | 164 (8.8%) | 145 (9.2%) | 21 (6.0%) | 19 (10.2%) | 105 (10.1%) | 19 (6.5%) |
| Divorced/separated | 179 (9.6%) | 157 (10.0%) | 28 (8.0%) | 20 (10.8%) | 109 (10.5%) | 22 (7.5%) |
| Widowed | 342 (18.3%) | 327 (20.7%) | 88 (25.1%) | 20 (10.8%) | 219 (21.0%) | 15 (5.1%) |
| **Educational Attainment, n (%)** |  |  |  |  |  |  |
| 5th–8th grade | 524 (28%) | 404 (25.6%) | 3 (0.9%) | 6 (3.2%) | 395 (37.9%) | 120 (40.8%) |
| 9th–12th grade | 374 (20%) | 291 (18.5%) | 32 (9.1%) | 31 (16.7%) | 228 (21.9%) | 83 (28.2%) |
| Technical School | 72 (3.8%) | 50 (3.2%) | 6 (1.7%) | 7 (3.8%) | 37 (3.6%) | 22 (7.5%) |
| Some University | 184 (9.8%) | 167 (10.6%) | 31 (8.9%) | 15 (8.1%) | 121 (11.6%) | 17 (5.8%) |
| University | 717 (38.3%) | 665 (42.2%) | 278 (79.4%) | 127 (68.3%) | 260 (25%) | 52 (17.7%) |
| **Currently Employed, , n (%)** | 415 (27.0%) | 292 (23.4%) | 50 (16.6%) | 46 (27.1%) | 196 (25.2%) | 123 (42.8%) |
| **Self-rated Health<sup>a</sup>, n (%)** |  |  |  |  |  |  |
| Excellent/very good | 238 (12.7%) | 210 (13.3%) | 87 (24.9%) | 35 (18.8%) | 88 (8.5%) | 28 (9.5%) |
| Good | 756 (40.4%) | 652 (41.4%) | 186 (53.1%) | 99 (53.2%) | 367 (35.3%) | 104 (35.4%) |
| Fair/moderate | 713 (38.1%) | 575 (36.5%) | 73 (20.9%) | 48 (25.8%) | 454 (43.7%) | 138 (46.9%) |
| Poor | 124 (6.6%) | 102 (6.5%) | 4 (1.1%) | 4 (2.2%) | 94 (9.0%) | 22 (7.5%) |
| Very poor | 39 (2.1%) | 37 (2.3%) | 0 (0.0%) | 0 (0.0%) | 37 (3.6%) | 2 (0.7%) |
| **BMI, kg/m^2, mean (SD)** | 29.07 ± 5.67 | 29.03 ±5.81 | 27.50 ±4.52 | 27.98± 4.92 | 29.71 ±6.19 | 29.27 ±4.92 |
| **Hypertension, n (%)** |  |  |  |  |  |  |
| No | 963 (51.5%) | 804 (51.0%) | 198 (56.6%) | 109 (58.6%) | 497 (47.7%) | 159 (54.1%) |
| Yes | 908 (48.5%) | 773 (49.0%) | 152 (43.4%) | 77 (41.4%) | 544 (52.3%) | 135 (45.9%) |
| **Diabetes<sup>a</sup>, n (%)** |  |  |  |  |  |  |
| No | 1430 (76.5%) | 1229 (78.0%) | 298 (85.1%) | 159 (85.5%) | 772 (74.2%) | 201 (68.4%) |
| Yes | 440 (23.5%) | 347 (22.0%) | 52 (14.9%) | 27 (14.5%) | 268 (25.8%) | 93 (31.6%) |
| **Hypercholesterolemia<sup>a</sup>, n (%)** |  |  |  |  |  |  |
| No | 883 (47.3%) | 715 (45.4%) | 89 (47.8%) | 173 (49.4%) | 453 (43.6%) | 168 (57.3%) |
| Yes | 985 (52.7%) | 860 (54.6%) | 97 (52.2%) | 177 (50.6%) | 586 (56.4%) | 125 (42.7%) |
| **Depressive Symptoms, mean (SD)** | 8.69 ± 6.04 | 8.70 ± 6.02 | 5.75 ± 4.60 | 6.13 ± 4.58 | 10.16 ± 6.16 | 8.64 ± 6.13 |
| **Physical Activity (IPAQ)<sup>a</sup>, n (%)** |  |  |  |  |  |  |
| Vigorous | 198 (10.6%) | 160 (10.1%) | 55 (15.7%) | 33 (17.7%) | 72 (6.9%) | 38 (12.9%) |
| Moderate | 1060 (56.7%) | 951 (60.3%) | 182 (52.0%) | 107 (57.5%) | 662 (63.7%) | 109 (37.1%) |
| Walking | 1757 (94.2%) | 1488 (94.7%) | 325 (92.9%) | 177 (95.2%) | 986 (95.2%) | 269 (91.5%) |
| None | 58 (3.1%) | 41 (2.6%) | 15 (4.3%) | 4 (2.2%) | 22 (2.1%) | 17 (5.8%) |
| **Currently Smoking, n (%)** |  |  |  |  |  |  |
| No | 1299 (69.6%) | 1110 (70.6%) | 145 (78.4%) | 300 (86.0%) | 665 (64.0%) | 189 (64.5%) |
| Yes | 567 (30.4%) | 463 (29.4%) | 40 (21.6%) | 49 (14.0%) | 374 (36.0%) | 104 (35.5%) |

<sup>a</sup>Missing/non-response; self-rated health, self-reported diabetes, and moderate activity, n = 1 each in the Total sample, Total matched sample, and CR-matched; self-reported hypercholesterolemia, n = 3 in the Total sample, 2 in each of the Total matched sample and CR-matched, and 1 in CR Supplemental; ever smoked during lifetime, n = 5 in the Total sample, 4 in the Total matched sample, 1 in UfSP, 1 in UfSN, 2 in CR-matched, and 1 in CR Supplemental; currently smokes cigarettes, n = 984 in the Total sample, 847 in the Total matched sample, 219 in UfSP, 104 in UfSN, 524 in CR-matched, and 137 in CR Supplemental; walking, n = 5 each in the Total sample, Total matched sample, and CR-matched; and any physical activity, n = 3 each in the Total sample, Total matched sample, and CR-matched.

### 3.4 Late Life Learning Exposure Characteristics Among Previously Enrolled UfS Participants

Characteristics of LLL participation among previously enrolled UfS participants are summarized in **Table 5**. On average, participants attended 7.0 UfS terms (SD=6.8) over a period of 4.82 years (SD=4.20). The most commonly reported reason for joining UfS, for which multiple responses were permitted, was a desire to learn more (51.1%), followed by socializing and meeting new people (33.1%), filling free time (32.0%), and a combination of other reasons (32.6%).

**Table 5.** Characteristics of UfS participation history among enrolled participants (N=350), Late Life Learning, Cognition and Aging Study (3LC), Beirut, Lebanon, 2025-2026.

| UfS characteristic | Mean (SD) or n(%) |
| --- | --- |
| **UfS Participation History** |  |
| Number of UfS terms attended | 7 ± 6.8 |
| Duration of participation(years) | 4.82 ± 4.20 |
| Cumulative participation time (years) | 3.51 ± 3.39 |
| **Reasons for joining UfS<sup>a</sup>** |  |
| Wanted to learn more | 179 (51.1%) |
| To break loneliness or isolation | 21 (6.0%) |
| For socializing or meeting new people | 116 (33.1%) |
| To fill free time | 112 (32.0%) |
| Encouragement from friends or colleagues in the program | 70 (20.0%) |
| To be associated with AUB | 41 (11.7%) |
| Other | 114 (32.6%) |
| **UfS activities participated in<sup>a,b</sup>** |  |
| Attending lectures | 340 (98.3%) |
| Participating in study groups | 227 (65.6%) |
| Participating in social activities | 235 (67.9%) |
| Participating in educational trips | 182 (52.6%) |
| **Change in attendance across terms<sup>b,c</sup>** |  |
| Participation increased across terms | 54 (20.1%) |
| Remained more or less the same | 114 (43.2%) |
| Participation decreased across terms | 97 (36.7%) |
| **Ever attended UfS activities online on Zoom<sup>b,c</sup>** |  |
| No | 29 (11.5%) |
| Yes | 224 (88.5%) |
| **Influence of Zoom options on attendance<sup>b,c</sup>** |  |
| Positively influenced attendance | 104 (46.2%) |
| Did not influence attendance | 71 (31.6%) |
| Negatively influenced attendance | 50 (22.2%) |
| **Preferred mode of delivery** |  |
| Almost all in person (more than 80%) | 113 (50.0%) |
| Mostly in person (>60%-80%) | 37 (16.4%) |
| Almost half in person (40%-60%) | 57 (25.2%) |
| Occasionally in person (>20%-40%) | 19 (8.4%) |
| Rarely in person (less than 20%) | 0 (0.0%) |

<sup>a</sup> Multiple responses were permitted. <sup>b</sup> 4 participants missing responses for UfS activities participated in, 5 participants missing responses for change in attendance across terms, 3 participants missing responses for ever attending UfS activities on zoom, and 1 participant missing response for influence of zoom options on attendance. <sup>c</sup> 80 participants were not eligible to answer this question as they have attended less than two terms, 94 were not eligible to answer as this question was restricted to those who attended UfS terms 2020 and/or onwards, 124 were not eligible to answer the influence on zoom options on attendance and preferred mode of delivery.

Engagement extended across several UfS activities. Nearly all participants attended lectures (98.3%), while approximately two-thirds participated in social activities (67.9%) and study groups (65.6%), and just over half participated in educational trips (52.6%). Across participation terms, involvement remained relatively stable for most individuals (43.2%), although 36.7% reported decreased participation over time and 20.1% reported increased participation.

## 3. DISCUSSION

The 3LC cohort establishes a resourceful platform for addressing critical gaps in understanding the life course determinants of cognitive aging in Lebanon and the broader MENA region, with a particular focus on investigating how under-examined psychosocial pathways and potentially modifiable later-life factors, such as late life learning, may shape cognitive health and resilience. In this cohort profile paper, we describe the 3LC design and baseline data collection among 1,871 adults aged 50 years and older, and highlight key features of the cohort, including its LLL-focused sampling design and comprehensive characterization of psychosocial, socioeconomic, and conflict-related adversity across the life course as well as psychosocial resilience measures. Despite recruitment taking place amid substantial economic and political instability and periods of war, the study successfully completed enrolment and baseline data collection with high completion rates. Informal feedback received from participants highlighted their appreciation of the interview experience, the relevance and comprehensiveness of the questionnaire, and the professionalism of the research team.

A distinctive feature of the 3LC study is the integration of a long-standing LLL program within a longitudinal aging cohort. This was supported by the longstanding infrastructure of the UfS, which provided an established framework for recruiting previously enrolled UfS participants with heterogeneous histories of participation and newly enrolled participants assessed before initiating LLL. UfS participants were predominantly women, consistent with patterns observed in voluntary LLL programs internationally^52,53^. Recruitment of age-, sex-, and education-matched community participants without prior LLL exposure enhanced comparability between LLL participants and community residents, while supplemental community recruitment enriched enriching the demographic and socioeconomic composition of the cohort. Together, the established UfS and the research platform developed through 3LC demonstrate the feasibility of implementing a comprehensive longitudinal cognitive aging study in this setting. Baseline data demonstrate substantial variation in the duration and intensity of LLL engagement across educational and social activity, providing a foundation for examining multiple dimensions of LLL participation in relation to cognitive and health trajectories.

Several LLL programs for older adults have been established internationally, including the Harvard Institute for Learning in Retirement^54^, 55 Plus Program at the University of Winnipeg^55^, Late Life Learning Toronto^56^, University of the Third age^57^, the Seniors University of China^58^ and other university-based lifelong learning initiatives across Europe, Asia, and North America^54–62^. These programs have contributed important insights into the role of later-life educational engagement in promoting social participation, wellbeing, and quality of life among older adults. However, many of these programs were primarily developed as educational and social engagement initiatives rather than research cohorts designed to evaluate LLL and cognitive aging outcomes. Consequently, rigorous empirical evidence regarding their potential cognitive and health benefits and the mechanisms through which these benefits might arise remain limited.

A limited number of studies have specifically examined the relationship between later life education and cognitive outcomes. For instance, findings from the Tasmanian Health Brain Project, a non-randomized longitudinal study of 459 healthy adults aged 50-79, show that participants who engaged in later-life university demonstrated improvements in language processing and verbal learning, contributing to enhanced cognitive reserve over four and seven years^63,64^. Similarly, in a longitudinal analysis of 12,099 adults aged 65 years and older from the Health and Retirement Study, participation in self-reported later-life learning activities was positively associated with cognitive function over time^23^. However, existing evidence has been derived from high-income settings and populations with different educational, social, and historical contexts. The 3LC cohort extends this evidence by assessing an LLL program within an LMIC setting and integrating detailed measures of cognition, social, biological, and seriocomic factors, and life-course exposures.

The study is thus uniquely positioned to investigate whether LLL can support cognitive aging and health in middle and older age, and whether it may buffer the effects of accumulated life-course adversity. These questions are particularly relevant in Lebanon and similar LMIC and resource-constrained settings, where opportunities for cognitive, social, and physical engagement in later life may be limited, while exposure to cardiometabolic and psychosocial risk factors for dementia is substantial. As a potentially scalable and modifiable strategy that can be implemented in later life, LLL could represent an important opportunity for promoting cognitive resilience and dementia prevention, including among individuals with substantial earlier-life or cumulative adversity. 3LC provides a unique platform to evaluate this potential and to determine whether, how, and for whom LLL may contribute to healthier cognitive aging, generating evidence to inform its potential as a scalable prevention strategy in diverse settings globally.

The scientific scope of 3LC extends beyond the evaluation of LLL. The cohort provides a platform for investigating cognitive aging in a population exposed to an unusual concentration of social, economic, and political adversity across the life course. Its comprehensive characterization of exposures including war and conflict, socioeconomic instability, migration and family separation, social relationships, and resilience, alongside cognitive, health, sensory, and biological measures, enables investigation of pathways through which cumulative adversity may shape dementia risk and of factors that may promote resilience. Together, these features position 3LC to contribute evidence on both risk and resilience from a population that remains markedly underrepresented in global cognitive aging and dementia research.

The design of the 3LC cohort includes some methodological considerations that should be acknowledged. First, although the study covers a large region within Beirut, participants enrolled in 3LC may not be generalizable to all older adults in Lebanon. Second, participants exposed to UfS may differ from community residents on characteristics other than age, sex and education (which are our matching criteria), such as health behaviors and socioeconomic factors. Therefore, future analyses examining the relationship between LLL and cognitive outcomes will need to carefully consider the effects of selection into the UfS.

Overall, the 3LC study provides a strong foundation for future longitudinal investigations of life course determinants of cognitive health and well-being in middle-aged and older adults in Lebanon. Through the development of comprehensive survey and collection of multidimensional data, 3LC represents an invaluable resource for examining not only risk factors for cognitive decline but also potential pathways underlying cognitive resilience, especially those that relate to late life learning.

## Data Availability

Data generated in this study are available from the authors upon request.

## FUNDING & ACKNOWLEDGMENTS

The 3LC study was funded by the National Institutes of Health, National Institute on Aging R01AG074076. We would like to thank 3LC study participants.

